# A novel specific artificial intelligence-based method to identify COVID-19 cases using simple blood exams

**DOI:** 10.1101/2020.04.10.20061036

**Authors:** Felipe Soares

## Abstract

**Background:** The SARS-CoV-2 virus responsible for COVID-19 poses a significant challenge to healthcare systems worldwide. Despite governmental initiatives aimed at containing the spread of the disease, several countries are experiencing unmanageable increases in the demand for ICU beds, medical equipment, and larger testing capacity. Efficient COVID-19 diagnosis enables healthcare systems to provide better care for patients while protecting caregivers from the disease. However, many countries are constrained by the limited amount of test kits available, lack of equipment and trained professionals. In the case of patients visiting emergency rooms (ERs) with a suspect of COVID-19, prompt diagnosis may improve the outcome and even provide information for efficient hospital management. In such a context, a quick, inexpensive and readily available test to perform an initial triage in ERs could help to smooth patient flow, provide better patient care, and reduce the backlog of exams.

**Methods:** In this Case-control quantitative study, we developed a strategy backed by artificial intelligence to perform an initial screening of suspect COVID-19 patients. We developed a machine learning classifier that takes widely available simple blood exams as input and classifies samples as likely to be positive (having SARS-CoV-2) or negative (not having SARS-CoV-2). Based on this initial classification, positive cases can be referred for further highly sensitive testing (e.g. CT scan, or specific antibodies). We used publicly available data from the Albert Einstein Hospital in Brazil from 5,644 patients. Focusing on simple blood exam figures as main predictors, a sample of 599 subjects that had the fewest missing values for 16 common exams were selected. From these 599 patients, 81 tested positive for SARS-CoV-2 (determined by RT-PCR). Based on the reduced dataset, we built an artificial intelligence classification framework, ER-CoV, aiming at determining if suspect patients arriving in ER were likely to be negative for SARS-CoV-2, that is, to predict if that suspect patient is negative for COVID-19. The primary goal of this investigation is to develop a classifier with high specificity and high negative predictive values, with reasonable sensitivity.

**Findings:** We identified that our AI framework achieved an average specificity of 85.98% [95%CI: 84.94 – 86.84] and negative predictive value (NPV) of 94.92% [95%CI: 94.37% – 95.37%]. Those values are completely aligned with our goal of providing an effective low-cost system to triage suspect patients in ERs. As for sensitivity, our model achieved an average of 70.25% [95%CI: 66.57% – 73.12%] and positive predictive value (PPV) of 44.96% [95%CI: 43.15% – 46.87%]. The area under the curve (AUC) of the receiver operating characteristic (ROC) was 86.78% [95%CI: 85.65% – 87.90%]. An error analysis (inspection of which patients were misclassified) identified that, on average, 28% of the false negative results would have been hospitalized anyway; thus the model is making mistakes for severe cases that would not be overlooked, partially mitigating the fact that the test is not highly sensitive. All code for our AI model, called ER-CoV is publicly available at https://github.com/soares-f/ER-CoV.

**Interpretation:** Based on the capacity of our model to accurately predict which cases are negative from suspect patients arriving in emergency rooms, we envision that this framework may play an important role in patient triage. Probably the most important outcome is related to testing availability, which at this point is extremely low in many countries. Considering the achieved specificity, we could reduce by at least 90% the number of SARS-CoV-2 tests performed in emergency rooms, with around 5% chance of getting a false negative. The second important outcome is related to patient management in hospitals. Patients predicted as positive by our framework could be immediately separated from other patients while waiting for the results of confirmatory tests. This could reduce the spread rate within hospitals since in many of them all suspect cases are kept in the same ward. In Brazil, where the data was collected, rate infection is starting to quickly spread and the lead time of a SARS-CoV-2 may be up to 2 weeks.

## Introduction

The SARS-CoV-2 virus responsible for Covid19 has posed a significant challenge to healthcare systems worldwide ^1^. Despite governmental initiatives aimed at containing the spread of the disease, several countries are experiencing unmanageable increases in the demand for ICU beds, medical equipment, and larger testing capacity. By April 5, 2020, more than 1.2 million people were infected by the new coronavirus, with over 60,000 deaths according to the World Health Organization (WHO) Situation Report 76^a^.

Efficient COVID-19 diagnosis enables healthcare systems to provide better care to patients while protecting caretakers from the disease. Most tests for the SARS-CoV-2 virus responsible for COVID-19 may either (i) detect the presence of a virus or compounds related to it, called molecular test, or (ii) detect antibodies produced as a reaction for virus exposure ^2^. Tests of type (i) are usually related to the Polymerase Chain Reaction (PCR), being labor-intensive in terms of laboratory procedures ^3^. Tests of type (ii) usually detect the IgG and IgM immunoglobulins related to SARS-CoV-2 and can be commercialized in the form of rapid tests ^2^. The global clinical industry is currently incapable of meeting the demand for SARS-CoV-2 tests. For instance, in Brazil, with a population of more than 200 million people, only 500,000 rapid tests were available at the end of March/2020, according to the Health Ministry^b^.

The frontline of medical care for COVID-19 is the emergency rooms at hospitals or health centers, which have to identify patients with COVID-19 from others with similar symptoms, such as fever, cough, dyspnoea, and fatigue, caused by other respiratory diseases ^4^. The quick determination of patient status regarding COVID-19 may determine follow-up procedures that may improve the overall patient outcome, as well as protect medical professionals. Thus, quick, inexpensive and broadly available proxy tests for COVID-19 are highly desirable.

## Research in context

### Evidence before this study

We searched PubMed on April 7, 2020, for studies using the terms (“SARS-CoV-2” OR “COVID-19” OR “coronavirus”) AND (“artificial intelligence” or “machine learning” or “data science”) without any restrictions regarding language or article type. We found no articles describing predictive methods (tests) for suspected patients based only on blood components. Articles reporting the use of artificial intelligence for COVID-19 diagnosis were found in the context of automatic assessment of chest CT ^5^. Previous studies correlated the C reactive protein to viral infections, mildly elevated alanine aminotransferase (ALT) in acute respiratory distress syndrome (ARDS) patients, leukopenia and lymphopenia ^6^.

A recent literature review on prediction models for diagnosis and prognosis of COVID-19 infection ^7^ listed 19 different models, including predictors such as sex, comorbidities, C-reactive protein, lymphocyte markers (percentage or neutrophil-to-lymphocyte ratio), lactate dehydrogenase, and features derived from CT images. None of the models predicted infection from blood exams data.

### Added value of this study

This is the first study to report a predictive system with high specificity and acceptable sensibility for the triage of suspected cases of COVID-19 in emergency rooms using only simple blood exams. We achieved an average specificity of 85.98% with a negative predictive value of 94.92%. The average percentage of false negatives is only 4.03%.

More specific tests are already available in the market, but at the current moment of the SARS-CoV-2 pandemic, stocks are short and result delivery time is long. With our solution, practically any clinical laboratory would be able to produce the information that is used as input in our method (ER-CoV) at a very low cost.

### Implications of all the available evidence

Our findings support that it would be possible to reduce at least 86% the number of SARS-CoV-2 tests performed in emergency rooms, with the chance of getting a false negative at around 5%, thus making screening more accessible for the population. Secondary implications are related to patient management, since with daily blood samples it would be possible to track patients that may have developed COVID-19 inside the hospital, allowing the adoption of more efficient isolation measures.

Artificial Intelligence (AI) methods have already been used in other medical applications, such as for the detection of colorectal cancer using blood plasma ^8^, prediction of drug-plasma binding ^9^, and identification of patients with atrial fibrillation during sinus rhythm ^10^. In the field of metabolomics, AI also plays an important role ^11^.

Considering the aforementioned successful research integrating AI and Medicine, we propose ER-CoV, an artificial intelligence-based screening method that uses blood exams to triage patients suspect of COVID-19 arriving in emergency rooms.

## Methods

### Study population

We performed a cross-sectional study with a control group on data collected by the Brazilian Albert Einstein Hospital, a leading facility in the management of COVID-19 in the state of São Paulo. The data is of public access^c^ and anonymized, encompassing 5,644 patients, and 108 laboratory and medical exam outcomes.

According to the data provider, the anonymized data was collected from patients seen at the hospital’s emergency room who were initially suspect cases of COVID-19 according to the hospital’s workflow for SARS-CoV2^d^. The main criteria for a case to be considered as suspect are the same provided by WHO for global surveillance of COVID-19^e^.

From the 108 outcome variables available in the dataset, we focused our research on those that could be quickly obtained at the ER or from patient records for retrospective analysis; namely:

1. Hemogram:
  a. Red blood cells
  b. Mean corpuscular volume (MCV)
  c. Mean corpuscular hemoglobin concentration (MCHC)
  d. Mean corpuscular hemoglobin (MCH)
  e. Red blood cell distribution width (RDW)
  f. Leukocytes
  g. Basophils
  h. Monocytes
  i. Lymphocytes
  j. Platelets
  k. Mean platelet volume
2. Creatinine
3. Potassium
4. Sodium
5. C reactive protein
6. Age

The reference method for determining if the individual was positive for SARS-CoV-2 was reported to be the reverse-transcriptase polymerase chain reaction (RT-PCR) method, as described in the national Brazilian guidelines^f^.

### Proposed workflow for patient screening using artificial intelligence

We developed our solution for patients arriving at a hospital’s emergency room. In Figure 1 we depict the workflow for using the proposed Artificial Intelligence (AI) model.

**Figure 1:**
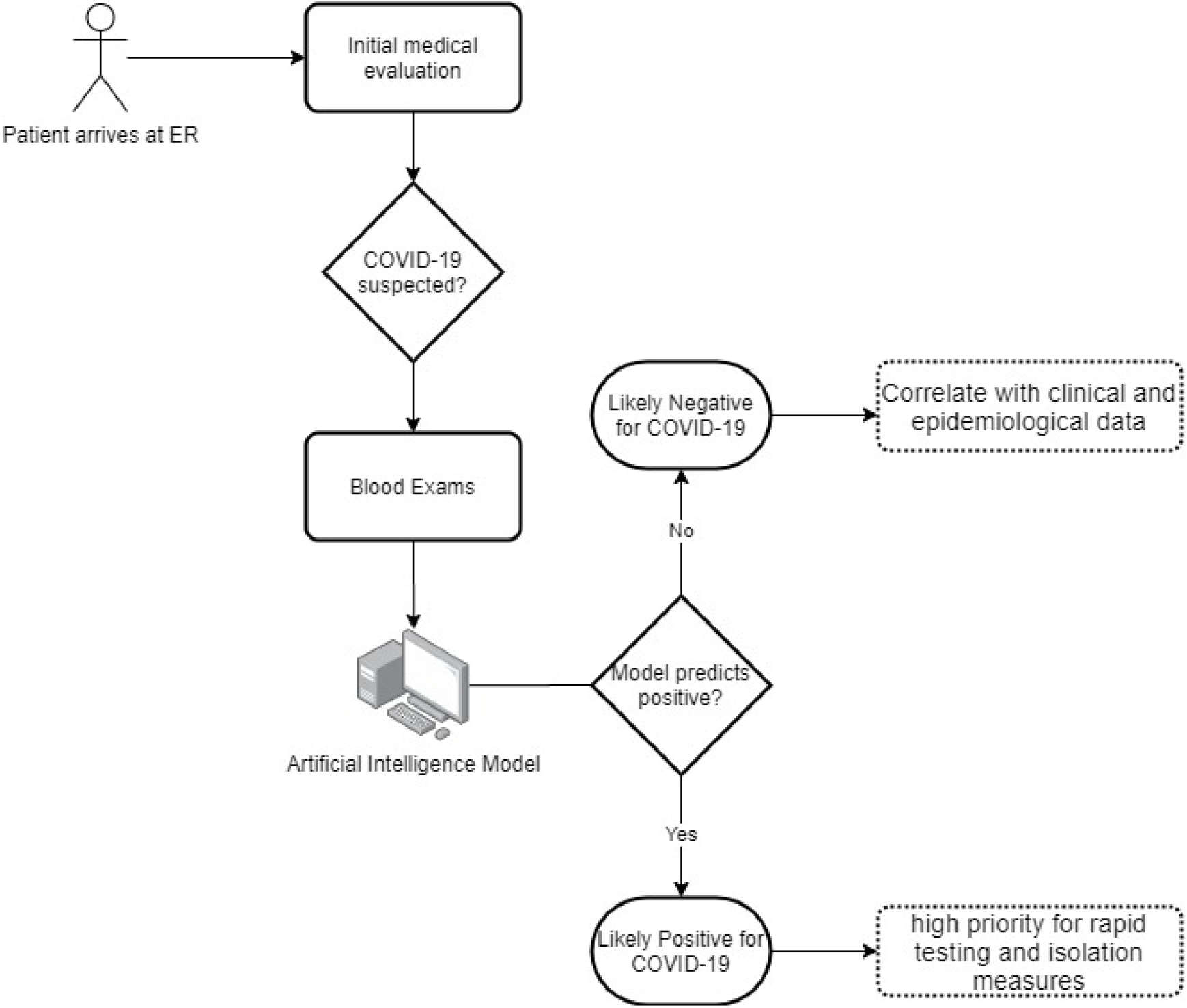
Diagram of the envisioned workflow using the ER-CoV artificial intelligence model in a hospital emergency room (ER).

A patient would initially be evaluated by health professionals to determine if she/he is a probable case of COVID-19. Once the patient is determined as a suspect case, simple blood exams would be requested, since they are the main input in the ER-CoV artificial intelligence model (see the previous section). Upon data entry (blood exams and age), the ER-CoV model yields a positive or negative result. In case of a positive result, considering the model’s relatively low sensitivity, priority should be given to this patient for further investigation including confirmatory PCR or antibody-based test for SSARS-CoV-2 and CT scan. If the result is negative, the patient is very likely not to have COVID-19.

### Proposed AI method for COVID-19 triage in ERs (ER-CoV)

The AI method uses 16 input variables and uses advanced techniques to predict the status of new patients regarding COVID-19. The flowchart for the proposed ER-Cov method is shown in Figure 2.

**Figure 2:**
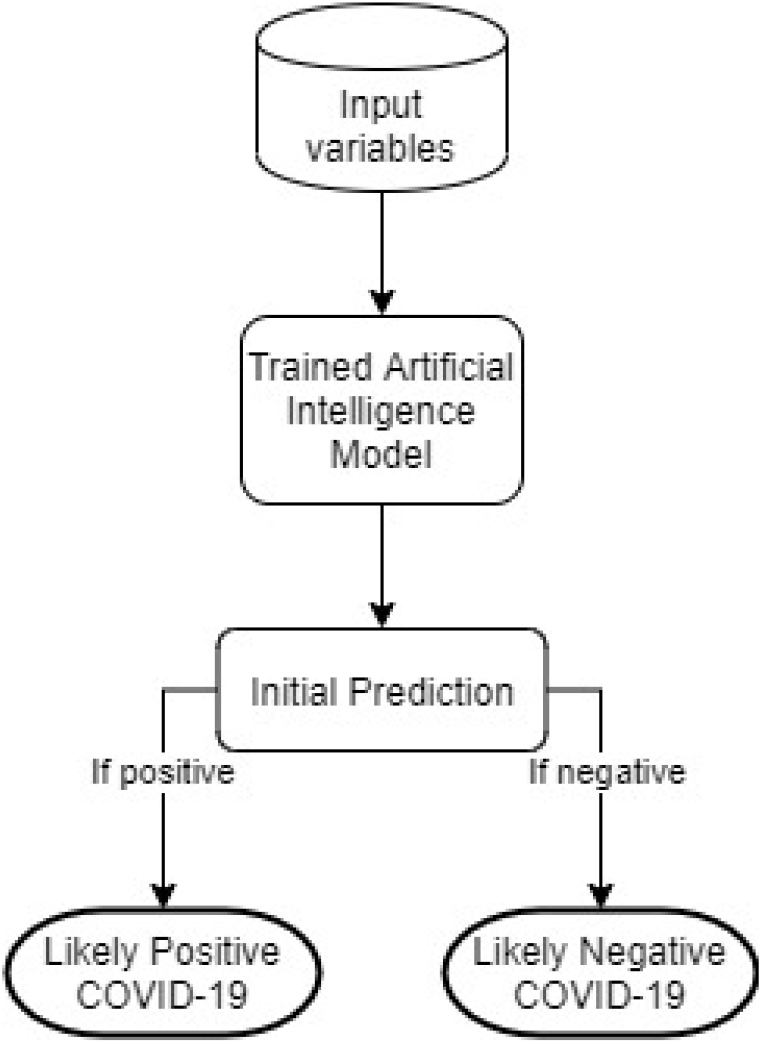
Flowchart for ER-CoV method for prediction of COVID-19 in suspect patients in ERs.

Once input variables are introduced in the ER-CoV model, an initial prediction of the ER patient’s COVID-19 status will become available

The AI model is trained using a combination of three techniques: Support Vector Machines ^12^, SMOTEBoost ^13^, and ensembling ^14^. Before training our model, C reactive protein missing values (present in 99 of the 599 samples in the reduced dataset) were imputed using the kNN algorithm with parameter *k* set to 5 ^15^.

Support Vector Machines (SVM) ^12^ is an AI technique used for binary classification (i.e. a given sample is classified as “positive” or “negative”). SVM has been successfully used in many biomedical applications ^8,16^. However, due to the small number of “positive” samples in the dataset (13.52%), the technique is prone to favor the majority class predicting all samples as “negative” and leading to extremely low sensitivity. To overcome such imbalance in classes, oversampling and ensemble methods were applied.

Oversampling was performed through the SMOTEBoost technique. SMOTE (Synthetic Minority Oversampling Technique) takes account of the relatively small number of positive samples by creating additional synthetic examples of this class. The boosting algorithm identifies incorrect predictions and assigns a larger importance weight for those samples, such that at the next iteration the algorithm is likely to display a smaller number of incorrect classifications. SMOTEBoost combines both approaches to improve classification performance for the imbalanced class. In addition, ensemble methods are based on the assumption that combining a collection of predictions from different models the final prediction will display better performance.

To develop the ER-CoV model we trained 10 SVM-based SMOTEBoost models. Its initial prediction corresponds to the average probability from all 10 models: if that probability is greater than 0.4, since we aim at taking a conservative threshold to minimize false negatives, the sample is classified as “positive”; otherwise, the model predicts the sample as “negative”. Details about model training are provided in the supplementary material, together with the source code. Incremental developments will be updated at https://github.com/soares-f/ER-CoV.

### Numerical Analysis

For the statistical analysis, we computed the following statistics: Sensitivity, Specificity, Positive Predictive Value (PPV), Negative Predictive Value (NPV), and the area under the curve (AUC) of the receiver operating characteristic (ROC) ^17^.

AI models may be affected by a specific partition of the data, that is, the particular samples used to calibrate and evaluate the model (i.e. training and test sets). To counter that, we repeated the process of training and evaluation 100 times using different partitions of data for training and test, and storing all information from each run. Approximately 90% of the data was assigned for training, and 10% for testing. Partitioning was carried out using stratified random sampling to ensure training and test sets with approximately the same proportion of positives and negatives.

If Influenza tests are not available, the sample will be classified as “likely positive” and submitted to SARS-CoV-2 specific tests. Analogously, if other rapid tests are available (e.g. for H1N1 Rhinovirus/Enterovirus), they could be added to the intermediary step for differential diagnosis before classifying the sample as “likely positive”.

After running these steps for ER-CoV, as depicted in Figure 2, we computed the average of the aforementioned statistics and their 95% confidence interval evaluated through BCa bootstrapping on the statistics collected from the 100 runs of ER-CoV with sample randomization ^18^. All models and statistics were obtained using R (version 3.6.3).

## Results

### Characterization of patients

A total of 599 patients were included in this study, corresponding to a subsample (“reduced dataset”) of the original dataset (“complete dataset”) of 5,644 individuals. The subsampling was performed to include only patients that had reported blood exams.

Due to the anonymization process performed by the hospital, we do not have access to the nominal values of the exams, but their standardized values (i.e. zero mean and unit standard deviation). The same is true for patient age, which was split in quantiles with no additional information given regarding the variable. Information on gender, race, and possible time of infection were not available.

There were 81 positive cases (13.52% prevalence) and 518 negative samples in the reduced dataset. The prevalence in the complete dataset, comprising 5,644 samples, was 9.88%. In Figure 3, we stratify the complete dataset into meaningful figures.

**Figure 3:**
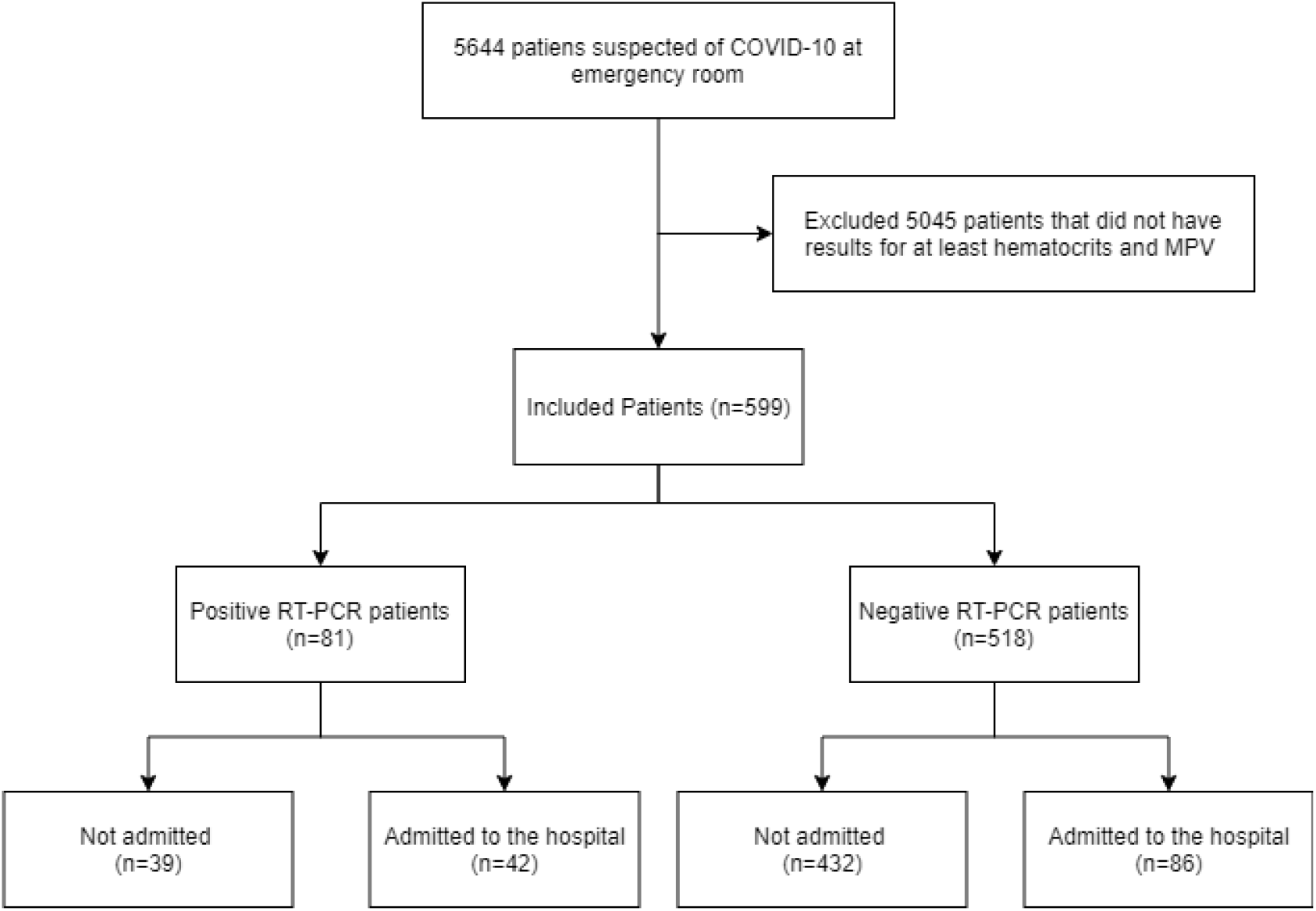
Diagram representing the process to select the included patients in our study.

### Results from the artificial intelligence predictive model

We applied the proposed AI predictive model to the reduced dataset. All results reported next were obtained from the 100 repetitions of training and testing, each repetition with a different sample distribution, thus minimizing the risk that our results are biased by a specific data partition. We included the confidence intervals reported for each metric.

The ER-CoV model’s specificity was 85.98% [95%CI: 84.94 – 86.84]; Sensitivity was 70.25% [95%CI: 66.57% – 73.12%] Negative Predictive Value (NPV) was 94.92% [95%CI: 94.37% – 95.37%]; Positive Predictive Value (PPV) was 44.96% [95%CI: 43.15% – 46.87%]. As for ROC AUC, ER-CoV achieved 86.78% [95%CI: 85.65% – 87.90%]. In Figures 4 to 7, we present the distribution of these metrics. Note that sensitivity and PPV values display a balanced behavior around their means; that is not observed in the skewed-to-the-right distributions of specificity and NPV values, which may be justified by their larger means.

**Figure 4:**
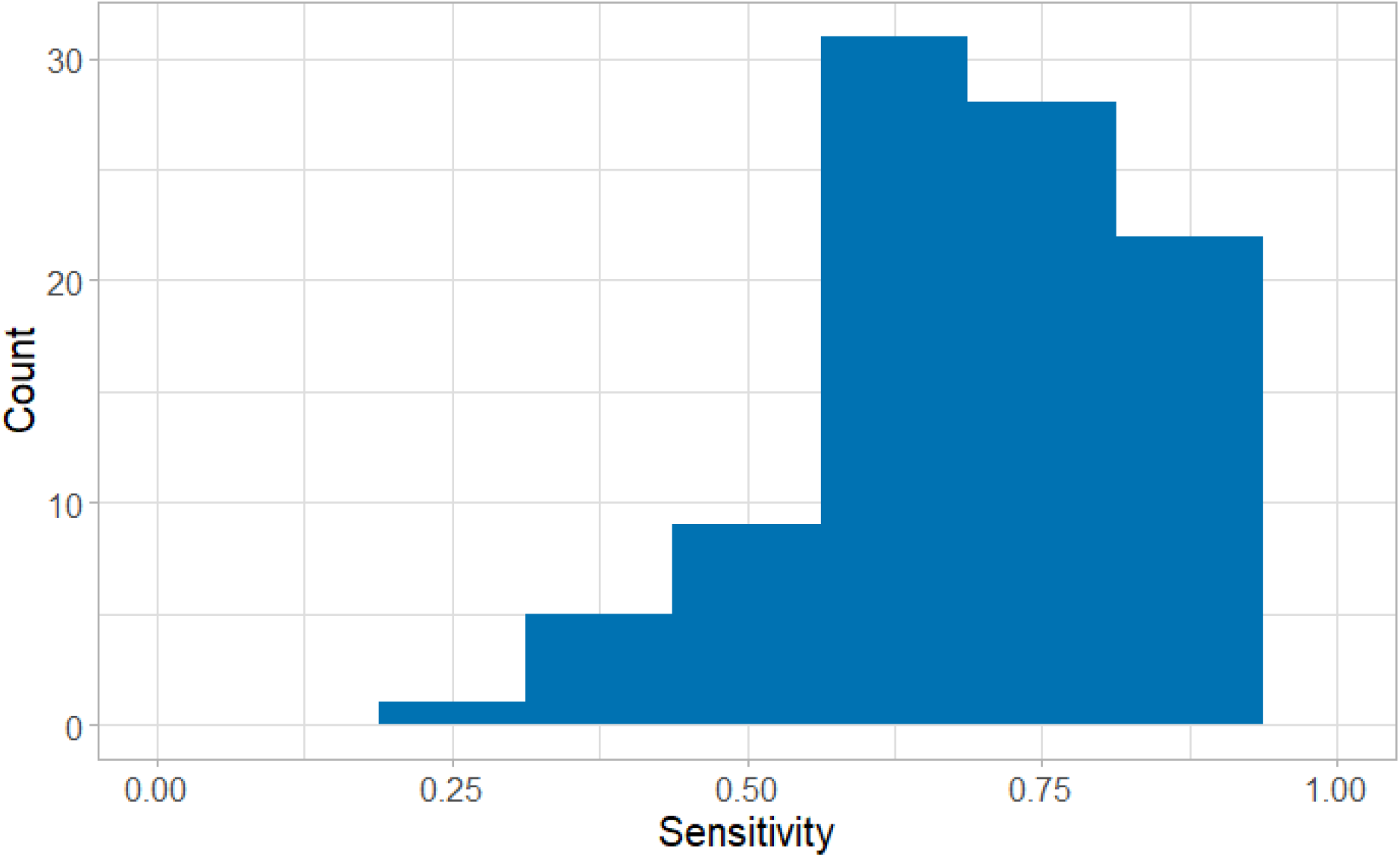
Histogram for the sensitivity of ER-CoV considering the 100 repetitions

**Figure 5:**
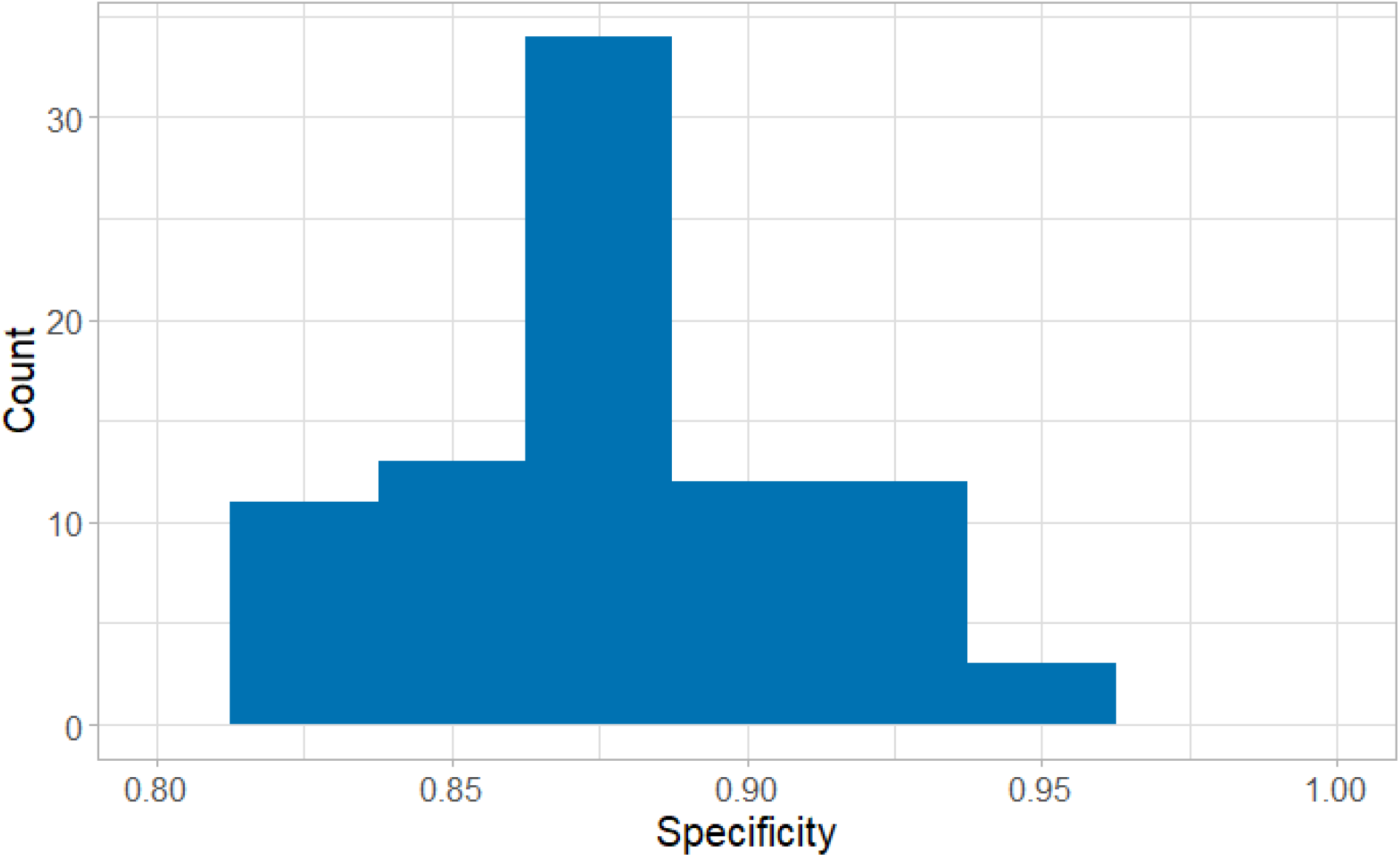
Histogram for the specificity of ER-CoV considering the 100 repetitions

**Figure 6:**
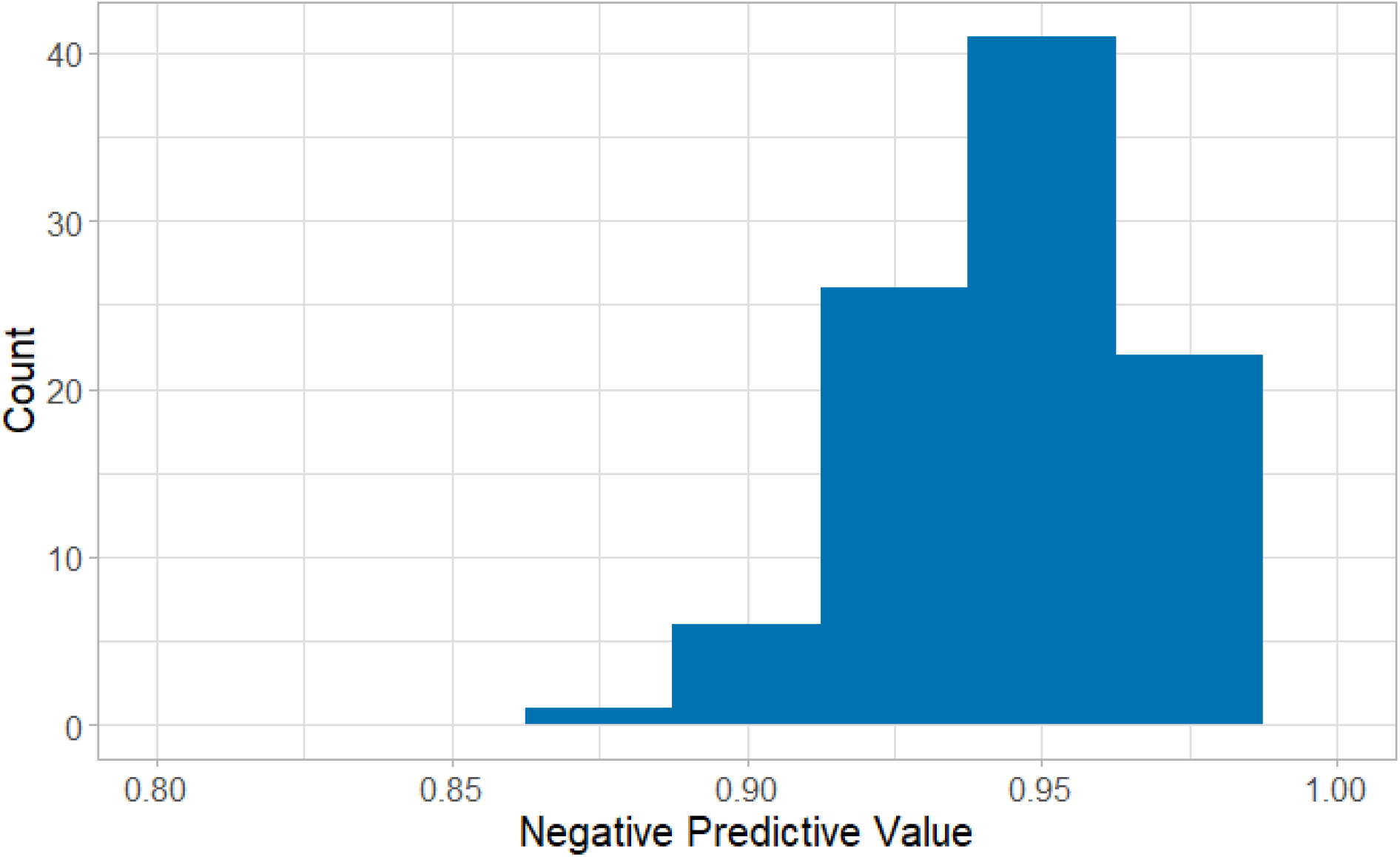
Histogram for the NPV of ER-CoV considering the 100 repetitions

**Figure 6:**
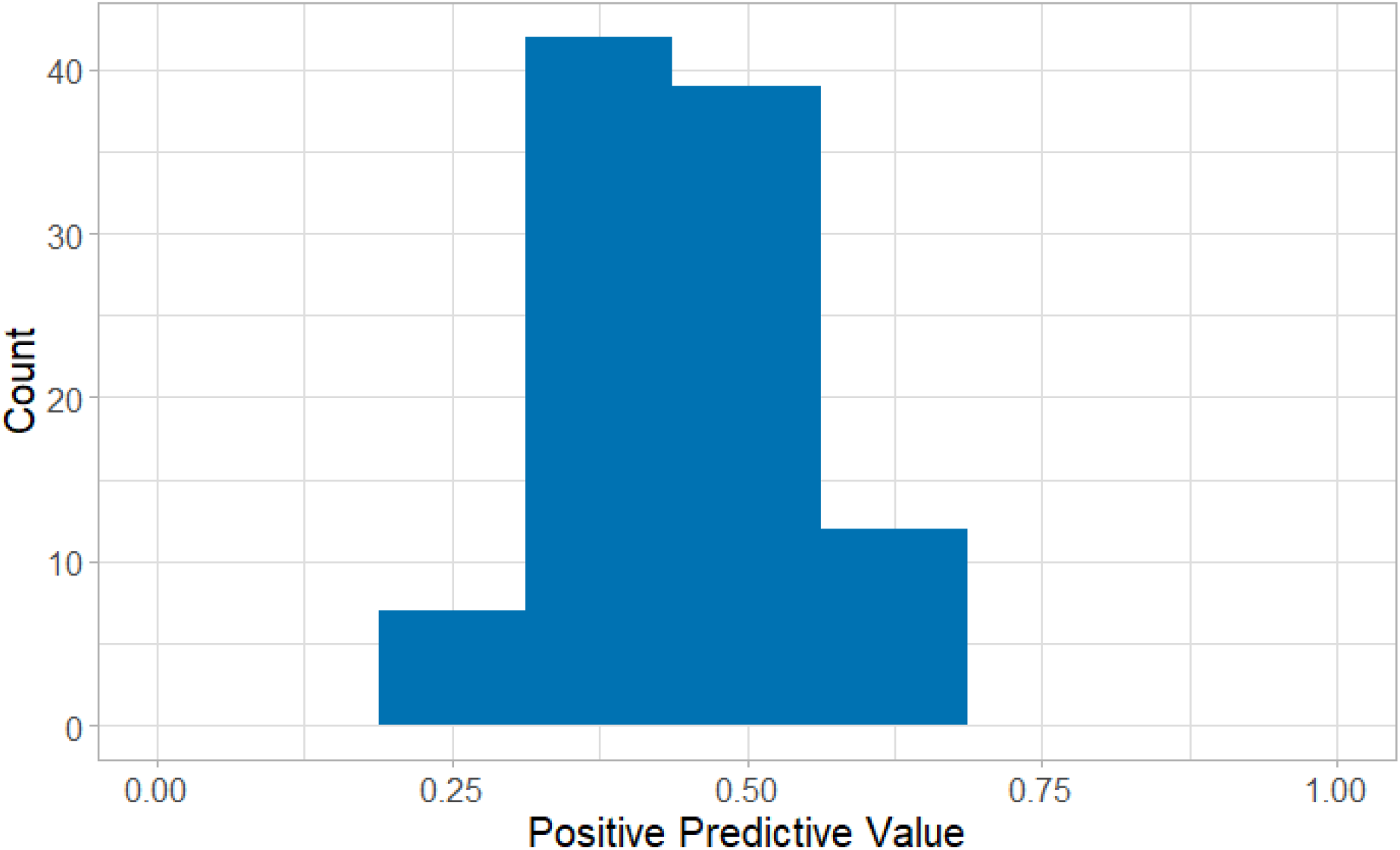
Histogram for the PPV of ER-CoV considering the 100 repetitions

Table 1 presents the average contingency table taken as percentages calculated over the 100 replications of our test. It is important to point to the low percentage of false negatives (4.03%), which corroborates the robustness of the ER-CoV model.

**Table 1:** Average contingency table obtained from 100 replications of ER-CoV

|  |  | Reference test (RT-PCR) |  |  |
| --- | --- | --- | --- | --- |
|  |  | Positive | Negative | Total |
| Our test (ER-CoV) | Positive | 9.53% | 12.88% | 22.41% |
|  | Negative | 4.03% | 73.56% | 77.59% |
|  |  | 13.56% | 86.44% | 100% |

We found that the proposed AI method was successful at discarding negative patients while flagging potential positive patients for COVID-19. When performing error analysis of the 100 replications, we found that 99 out of 599 patients were misclassified at least once, as shown in Table 2.

**Table 2:** Summary of the number of unique patients that were misclassified during the repetitions of our experiments and their status regarding hospital admission

|  | RT-PCR Positive | RT-PCR Negative |
| --- | --- | --- |
| Admitted | 9 | 16 |
| Non-Admitted | 23 | 97 |
| Total | 32 | 113 |

Of the patients misclassified as negative (false negatives), 28% were admitted to the hospital, possibly due to the severity of their symptoms or to other factors such as comorbidities. In comparison, only 9.31% of the COVID-19 positive patients in the complete dataset were hospitalized. Thus, even with an average sensitivity of 70.25% obtained using a limited number of blood exam variables (which may already be viewed as an important finding), the combination of ER-CoV with clinical features may improve COVID-19 diagnosis and guarantee that people receive the adequate care.

In our dataset 196 patients were tested for Influenza A and B and 20 (10%) had positive results. There were no positive samples of both Influenza (A or B) and COVID-19. We did a post-hoc analysis including Influenza in the model, but it did not change the model’s predictive value (<1% change in specificity).

For shortness, additional results obtained using other setups, such as different training and test ratios and other classifiers, are included as supplementary material. All results can be verified using the provided files.

## Discussion

By the simple introduction of the AI model, we can potentially improve COVID-19 screening in emergency rooms. Given its NPV of 94.92%, we would be able to identify patients with very low risk for COVID-19., The use of this tool along with clinical evaluation could reduce the number of traditional tests already adopted at the hospital’s ER. Therefore, the first (and possibly most important) benefit is the possible reduction in the number of tests required to be performed on patients that are negative for COVID-19. Another benefit is the potential to develop prioritization queues for patients that our model classifies as likely positive, thus speeding up results for potentially infected patients. That is, with a specificity of 85.98%, suspected COVID-19 cases would be prioritized for further testing. In a global scenario of an extreme shortage of testing kits and laboratory supplies for PCR related exams, we foresee that our proposed method may have a great impact on patient diagnosis in ERs, especially in developing and low-income countries.

RT-PCR tests remain the gold-standard for COVID-19 diagnosis; however, it presents some drawbacks. High viral loads are detected in the upper and lower respiratory tract after 5-6 days of symptoms onset, thus false negative results may be seen in the first days of infection. The sensibility of the test also depends on the site and technique used for swab collection. Repeating the exam may be necessary in suspected cases with initial negative results. Besides, processing of the sample is labor-intensive, and results may take from hours to few days to be available, depending on the setting. Rapid antigen lateral flow assays are point-of-care tests that have been developed with the advantage of fast results, but there is a concern about their sensitivity. Serologic tests are valuable for epidemiological purposes, but are best used retrospectively, once it takes weeks to develop specific IgG antibodies ^19^. Such time gap between the beginning of infection symptoms and definitive diagnosis is a challenge for managing patients with suspected COVID-19 infection. Having a model based on simple and fast biochemical laboratory exams may help hospitals to build a more efficient patient flow management, while definitive confirmatory exams are not available. Hospitals would be able to provide better isolation of COVID-19 patients, given that blood samples could be drawn on a daily basis and ER-CoV would identify patients that should be moved to another ward (say, a ward for suspected positive patients). Patients with positive model classification and negative PCR test outcomes in ER arrival could be closely followed and be candidates for repeating the test according to clinical evaluation.

Other rapid tests for viral infections (e.g. for Influenza A and B, H1N1, Rhinovirus/Enterovirus) could be added, when available, as an intermediary step for differential diagnosis before classifying the sample as “likely positive”. Co-infection of COVID-19 and Influenza is uncommon and has been reported in very few patients ^20^. In our dataset, no co-infection was reported among the 196 patients tested for both diseases. Unfortunately, most patients were not tested for Influenza and we could not report improvements in the model’s specificity by adding this variable to our analysis.

A limitation of our study is that we were not capable of identifying with high certainty which blood exams contribute the most to the classification, due to the nature of our AI model framework. However, previous studies already identified that C reactive protein ^21^, leukocytes ^22,23^, platelets ^22^, and lymphocytes ^24^ are altered at different levels in COVID-19 patients. Thus, we envision for future research a detailed study of which blood exams are more informative for differential diagnosis, as well as understanding how the SARS-CoV-2 virus alters blood components.

Another possible limitation is that data were collected only at the emergency room, with patients already displaying symptoms compatible with COVID-19. At this point, due to the lack of data from asymptomatic patients, we cannot generalize how our model would perform for a group of individuals that are not compatible with characteristic symptoms of COVID-19.

In this paper, we report a novel method for the classification of COVID-19 patients in emergency rooms. The ER-CoV method is low-cost and relies only on simple blood exams that are fast and highly available, and resort to artificial intelligence methods to model such patients. We achieved significant results and foresee many applications of this framework. We thus support additional initiatives such as this one by the Albert Einstein Hospital, since the availability of data on COVID-19 tests allows the proposition of AI-based methods that could support medical decision and may provide important insights in longitudinal studies on the progression of the disease.

## Data Availability

Source code is shared under AFPL License. No commercial derivatives are allowed without formal consent from the first author.

https://github.com/soares-f/ER-CoV

## Acknowledgments

This work was supported by the Amazon AWS Cloud Credits for Research, the Google TensorFlow Research Cloud for the free credits for the usage of TPU. Their role was to provide computational resources for all tested models.

The University of Sheffield had the role of financially supporting the Ph.D. scholarship of Felipe Soares.

Prof. Fogliatto’s research is funded by CNPq [Grant # 303509/2015-5].

Prof. Anzanello’s research is funded by CNPq [Grant # 306724/2018-9].

## Ethical Approval

We used only secondary anonymized data for our analyses. Self-declaration approval was given by the University of Sheffield Research Ethics Committee (Reference Number 034058).

## Footnotes

a https://www.who.int/docs/default-source/coronaviruse/situation-reports/20200405-sitrep-76covid-19.pdf?sfvrsn=6ecf0977_2

b https://www.saude.gov.br/noticias/agencia-saude/46623-brasil-inicia-a-distribuicao-de-500-miltestes-rapidos

c https://www.kaggle.com/einsteindata4u/covid19

d https://medicalsuite.einstein.br/pratica-medica/Documentos%20Doencas%20Epidemicas/Manejo-de-casos-suspeitos-de-sindrome-respiratoria-pelo-COVID-19.pdf - In Portuguese

e https://www.who.int/publications-detail/global-surveillance-for-human-infection-with-novel-coronavirus-(2019-ncov)

f https://portalarquivos2.saude.gov.br/images/pdf/2020/marco/04/2020-03-02-Boletim-Epidemiol--gico-04-corrigido.pdf

